# Leveraging proteomics and transfer learning for head and neck cancer detection in saliva

**DOI:** 10.1101/2025.07.13.25331474

**Authors:** Anza Shakeel, Samuel W. D. Merriel, Joel Smith, A. Stephen McGough, Matthew Suderman, Zahraa S. Abdallah, Paul D. Yousefi

**Affiliations:** MRC Integrative Epidemiology Unit, University of Bristol, Bristol, UK; NIHR Bristol Biomedical Research Center, University Hospitals Bristol and Weston NHS Foundation Trust and University of Bristol, Bristol, UK; College of Medicine and Health, University of Exeter, Street, Exeter, UK; School of Engineering Mathematics and Technology, University of Bristol, Street, Bristol, UK; School of Computing, Newcastle University, Newcastle, UK; Royal Devon and Exeter Foundation Trust, Exeter, UK

**Keywords:** Proteomics, Head and Neck Cancer, Deep Learning, Variational Autoencoder, Convolutional Neural Networks, Cancer Biomarkers

## Abstract

**Background:** Early detection of Head and neck cancer (HNC) has the potential to substantially improve patient survival, yet no biomarker tests for early detection are currently in clinical practice. Case-control studies that could be used to derive diagnostic biomarkers tend to be underpowered. Recent evidence suggests that we may be able to address this challenge by applying deep learning on pan-cancer data from large population studies.

**Methods:** We evaluate a range of machine learning methods and training scenarios to use proteome data to distinguish between HNC cases and controls. Models were trained on blood plasma proteomes from the UK Biobank (UKB) with n = 13,208 pan-cancer cases. To assess model’s generalisability across tissue types, we tested in a cross-tissue comparison using an independent saliva based proteome dataset from the SensOrPass HNC case-control study (n = 156).

**Findings:** We obtain best performance (AUC=0.88 versus AUC ***<*** 0.77 for others) using a transfer learning approach called CNN-Synth. This convolutional neural network was trained on UKB to distinguish between profiles from a set of controls and cases including synthetic profiles generated by a pretrained variational autoencoder. Post-hoc model explainability using SHapley Additive explanations identified IL6, CXCL17, CXCL13, IGF1R and FASLG as the top five proteins contributing most to predictor performance.

**Interpretation:** Our findings underscore the potential for deep learning and explainable AI to leverage data from large-scale population datasets for advancing early cancer detection and improving clinical outcomes.

**Funding:** This work was supported by Cancer Research UK (grant numbers EDDISA-Jan22\100003 and C18281/A29019).

## Research in context

### Evidence before this study

Survival in head and neck cancer (HNC) is dramatically improved when it is diagnosed early. Unfortunately, there are no screening programmes for HNC, so diagnosis tends to occur after the appearance of symptoms. Previous research has shown the potential of proteomic biomarkers as a screening tool for early cancer detection. Deep learning methods have shown promise for the discovery of biomarkers in high-dimensional data with potential to generalise beyond specific tissue types, cohorts, and cancer types and to overcome common challenges such as class imbalance, a frequent challenge for rare cancers like head and neck cancer (HNC).

### Added value of this study

This study introduces CNN-Synth, a deep learning model for estimating HNC risk from blood plasma protein abundance. CNN-Synth is a convolutional neural network (CNN) trained in UK Biobank (UKB) on a mix of real and synthetic pan-cancer proteomic profiles generated by a variational autoencoder (VAE) to reduce class imbalance. CNN-Synth is unique in that it did not require re-training or fine-tuning to accurately detect HNC cases in proteomes derived from saliva instead of blood. Furthermore, SNN-Synth outperformed classical machine learning models trained either from scratch in saliva proteomes or via transfer learning from blood proteomes.

### Implications of all the available evidence

These findings highlight the potential of generative deep learning approaches for the discovery of biomarkers with improved performances, particularly in low-sample or domain-shifted scenarios. CNN-Synth offers a scalable solution that can generalize across tissue types and cancer types without the need for fine-tuning. This supports broader use of representation learning in proteomics and suggests that models trained on large-scale plasma data can be effectively transferred to alternative biological matrices, opening new possibilities for biomarker discovery and detection in rare cancers.

## 1 Background

Head and neck cancer (HNC) incidence has been increasing globally and in the UK, where 58.5% of the time it is diagnosed at late stage, making it a major contributor to cancer-related mortality [1, 2]. Earlier detection could improve survival rates by identifying cancers when they are smaller, less invasive, and more treatable. Proteomic biomarkers offer a promising, non-invasive solution that are readily assayable in blood, saliva, or tissue samples, but their development is challenged by small case sizes, high-dimensional data leading to overfitting, and biological variability across cancers [3–6]. Population-scale protein data has already been observed to be a valuable source of information for cancer detection [3, 4, 7]. The UK Biobank’s (UKB) proteomic dataset, which includes 2, 941 proteins measured in plasma collected from 53,014 individuals, provides a valuable resource for identifying biomarkers and understanding disease mechanisms [8].

Machine learning approaches are well suited to extract disease-relevant signal from high-dimensional molecular datasets, including those with proteomic measures. Already, a wide range of machine learning techniques have been applied to predict disease risk and patient outcomes ([9–11]). Deep learning approaches, particularly convolutional neural networks (CNNs), have expanded predictive capabilities by learning hierarchical feature representations from high-dimensional data [12]. Although CNNs are best known for their applications in medical imaging, they are beginning to be applied to omic data for disease prediction [11, 13–15].

One very useful feature of deep learning models is their ability to generalize beyond the specific task for which they were trained. For example, CNNs trained in specific disease contexts have been found to capture variation relevant to multiple diseases, likely due to their capacity to characterize general biological relationships among molecular traits. This behavior has been leveraged, using an approach more generally called transfer learning [16], to use models trained for one outcome to generate models with improved performance for less common outcomes that lack sufficient training data [11, 17, 18]. Alternatively, another class of generative deep learning models, called Variational Autoencoders (VAEs), are specifically developed with the general aim of capturing low-dimensional representations of complex relationships between features. These models have been widely applied not only to generate synthetic data, for example, to address class imbalance in the context of rare cancers [11, 19, 20]—but also for dimensionality reduction and unsupervised clustering of high-dimensional biological data. In particular, VAEs have been used to uncover biologically meaningful protein modules from large-scale plasma proteomic datasets, as demonstrated by [15].

In this study, we apply VAE data simulation to address class imbalance and transfer learning across outcomes and tissue types for HNC detection. Our method is based on a CNN model pre-trained on blood plasma protein abundance data from a pan-cancer case-control study within a large, population cohort study which we then evaluate in saliva samples from a small HNC-specific case-control study. When training, we employ a VAE to generate synthetic cancer protein profiles to augment the number of cases. We show that the resulting model, CNN-Synth, performs better than classical machine learning approaches for distinguishing between HNC cases and controls. SHapley Additive exPlanations (SHAP) [21] for this model highlight a critical role for the well-known disease biomarker IL6.

## 2 Methods

### 2.1 Study populations

#### 2.1.1 UK Biobank (UKB)

UKB is a population-based cohort with data collected from over 500,000 adult participants aged between 40-69 years that were recruited between 2006 and 2010 in the UK. It includes extensive phenotypic, clinical, and multi-omic measurements. Our study used data from a subset of N = 54,219 UKB participants selected as part of the UKB Pharma Proteomics Project [8], hereafter referred to as UKB. These participants had 2,941 plasma protein analytes representing 2,923 unique proteins measured by the Olink Explore 3072 Protein Extension Assay (Uppsala, Sweden). Comprehensive details on proteomic data generation, along with normalization and quality control procedures, have been published [8, 22]. Protein levels were reported and analyzed as normalized protein expression (NPX) on the log base 2 scale. For compatibility with the measures available in our test set, we restricted all analysis to the 92 proteins that were also available on the Olink Oncology II panel used in the SensOrPass cohort.

Information on cancer incidence for UKB was obtained via the cancer register. We identified N = 13,208 cancer cases as those who had ‘reported occurrences of cancer’ and ‘date of cancer diagnosis’ for all instances in the cancer registry (Table 1 & Supplemental Table 3). The cases included a wide range of malignancies, including 14% with malignancies of the lip, oral cavity, pharynx, or oropharynx (Supplemental Figure 8). All other remaining participants were included as controls. Of these 13, 208 cases, n = 8, 262 where diagnosed after sample collection and n = 4, 945 where diagnosed before sample collection, the time-to-diagnosis distributions are shown in the Supplementary Figure 6. These counts are computed using the variable ‘Date of cancer diagnosis’ and ‘Sample collection sign-off timestamp’.

**Table 1.** Details of the SensOrPass study (saliva samples collected for this study) and UKB populations with proteome data.

| Study populations | Sample Size n | Age Mean (SD) | Disease Status | Case n(%) | Control n(%) | Tissue Type | Ethnicity |
| --- | --- | --- | --- | --- | --- | --- | --- |
| UK Biobank<br>(Training data) | 53,014 | 56.8<br>(8.2) | Pan-cancer | 13,208<br>(24.9%) | 39,806<br>(75.1%) | Plasma | ≈ 90% White British |
| SensOrPass<br>(Test data) | 156 | 63.8<br>(9.6) | Head and Neck Cancer | 64<br>(41.02%) | 92<br>(58.9%) | Saliva | 98.9% White British |

#### 2.1.2 SensOrPass head and neck cancer case-control study

SensOrPass is a N = 156 participant head and neck cancer (HNC) case-control study recruited from the south west of England with the aim of identifying saliva protein biomarkers of HNC cancer that would be suitable for passive monitoring with electrochemical sensors. N = 14 treatment-naive HNC cases were recruited at the Royal Devon University NHS Foundation Trust and Hampshire Hospitals NHS Foundation Trust. An additional N = 50 cases were selected from among the participants in the Head & Neck 5,000 cohort with cancer (stage I / II / III and IV) at diagnosis with available saliva samples (Figure 4 (c)) [23, 24]. A N = 92 control group was recruited from healthy volunteers of the Exeter 10,000 cohort [25]. The abundances of 92 proteins were measured in saliva samples using the Olink Oncology II panel (Table 1 & Supplemental Table 4).

### 2.2 Variational Autoencoder (VAE) of cancer-specific protein abundance

Toward the aim of simulating synthetic cancer-specific protein profiles, we trained a VAE from UKB cancer cases using the 92 Olink Explore 3072 proteins that were also available on the Oncology II platform in the SensOrPass validation data. The VAE architecture consisted of a protein input layer that is passed to an encoder with three fully connected layers of decreasing size–50, 25, and 10 neurons, respectively–to encode the input data into a lower-dimensional latent space (Figure 2(a)). Synthetic observations can be sampled from the encoder-generated latent space via a decoder of roughly symmetrical architecture, starting with a fully connected layer of size 10, followed by layers of 25 and 50 neurons and final output layer corresponding to the 92 reconstructed proteins. A Tanh activation function was used for every layer except the output layer. The model was optimized using the Adam optimizer and trained with a learning rate of 10*^−^*^6^.

To train the VAE, we defined a total VAE loss function to balance reconstruction accuracy with regularization of the latent space, allowing the model to learn meaningful representations of proteomic data while preventing overfitting. The total VAE loss is made up of two components: 1) the reconstruction loss and 2) the KL divergence loss.

For reconstruction loss, which quantifies the difference between the original input and its reconstruction by the decoder (eq: 1), we specified a mean squared error (MSE) loss function:

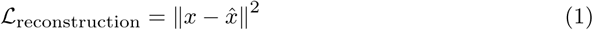

For KL divergence (eq: 2), we specified how much the learned latent distribution *q*(*z*|*x*) diverges from a standard normal prior distribution N (0*, I*):

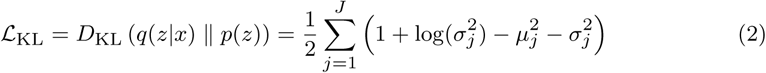

where *µ* and *σ* are the mean and standard deviation vectors from the encoder’s output, and *J* is the dimensionality of the latent space.

Total VAE loss combined both (eq: 1) and (eq: 2) as shown below:

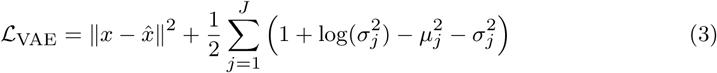

VAE training aimed to minimize (eq: 3) using all available UKB cancer cases (N = 13,208). Training progress was assessed by closely monitoring the training loss (Supplemental Figure 7(a)) and was halted when the reconstruction loss plateaued at 50 epochs.

### 2.3 Protein-based CNN cancer classifiers (CNN-Raw and CNN-Synth)

In UKB data, we trained two protein-based CNN cancer classification models [26, 27], CNN-Raw and CNN-Synth. Both models were trained using UKB plasma protein abundance data to distinguish between pan-cancer cases and controls. Training was identical for both models except that the CNN-Synth training dataset include 10,000 additional cancer-specific protein profiles synthesized by the VAE described above. These model inputs were presented as a 1×92 input vector of features, corresponding to those available in both UKB and SensOrPass, and the model output represented estimated probability of cancer.

We specified a network architecture consisting of two 1D convolutional layers, each using 64 filters with a kernel size of 1×1 (Figure 3). This enabled the extraction of high-level feature representations from the input vector, the spatial structure of which is inferred from the plasma proteomic data. Each convolutional layer *l* applied filters to the input through the following operation:

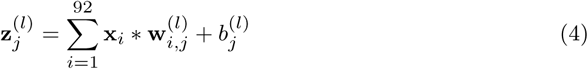

where *j* is the number of filters, **x_i_** is the input of each layer, **w**^(*l*)^ are the weights of the filter *j* and the input *i*, **b***_J_*^(*l*)^ is the bias for each filter, and ∗ denotes the 1D convolution operation. The length of the output feature map for each filter is computed as:

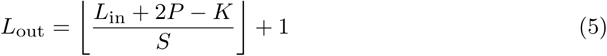

where *L*_in_ is the length of the input vector, *K* is the kernel size (1 in our case), *P* is the zero-padding size (typically 0 unless otherwise specified) and and *S* is the stride with which the kernel moves across the input vector. These convolutional layers are followed by a ReLU activation function before feeding into three fully connected layers intended to integrate and refine the extracted features for binary classification. The first and second fully connected layers contain 32 and 10 neurons, respectively. The final output is specified by a sigmoid function with output values between 0 (control) and 1 (case), which are interpreted as probabilities.

Training aimed to minimize the binary cross-entropy loss function (eq: 6), which measures how well the model’s predicted probability *p* aligns with the true label *y* [28]:

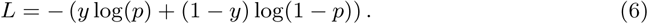

All models were trained using early stopping, where training was halted if the validation loss did not improve for a predefined number of consecutive epochs (i.e. 15), in order to prevent overfitting and ensure generalization. For the CNN-Synth model, early stopping occurred at epoch 50 (see Supplemental Figure 7(b)).

### 2.4 Classical protein-based ML cancer classifiers

To compare the performance of our protein-based generalized CNN cancer discrimination model against a wide range of alternative approaches, we trained a diverse library of classical machine learning methods for comparison to the CNN models described above (Section: 2.3). These span various algorithmic families with differing assumptions and inductive biases. Specifically, we evaluated logistic regression with L1/L2 regularization to mitigate overfitting and enhance generalization (LR-elasticnet), linear discriminant analysis (LDA), k-nearest neighbors (KNN), decision trees (CART), Gaussian naïve Bayes (NB), support vector machines (SVMs), and eXtreme Gradient Boosting (XGBoost), which are well-suited for handling high-dimensional and potentially sparse feature spaces. To ensure consistency and comparability, grid search with 10-fold cross validation was employed to tune the hyperparameters of each model, with the best hyperparameters selected based on accuracy. Classical ML model performances were evaluated in two different training-testing scenarios. In the first scenario, models were trained in UKB and tested in SensOrPass, similarly to the CNN models described above (transfer learning approach - left panel shown in Figure 1). In the second scenario, models were trained from scratch and performances evaluated within the context of 10-fold cross-validation using only SensOrPass data (right panel shown in Figure 1).

**Fig. 1.**
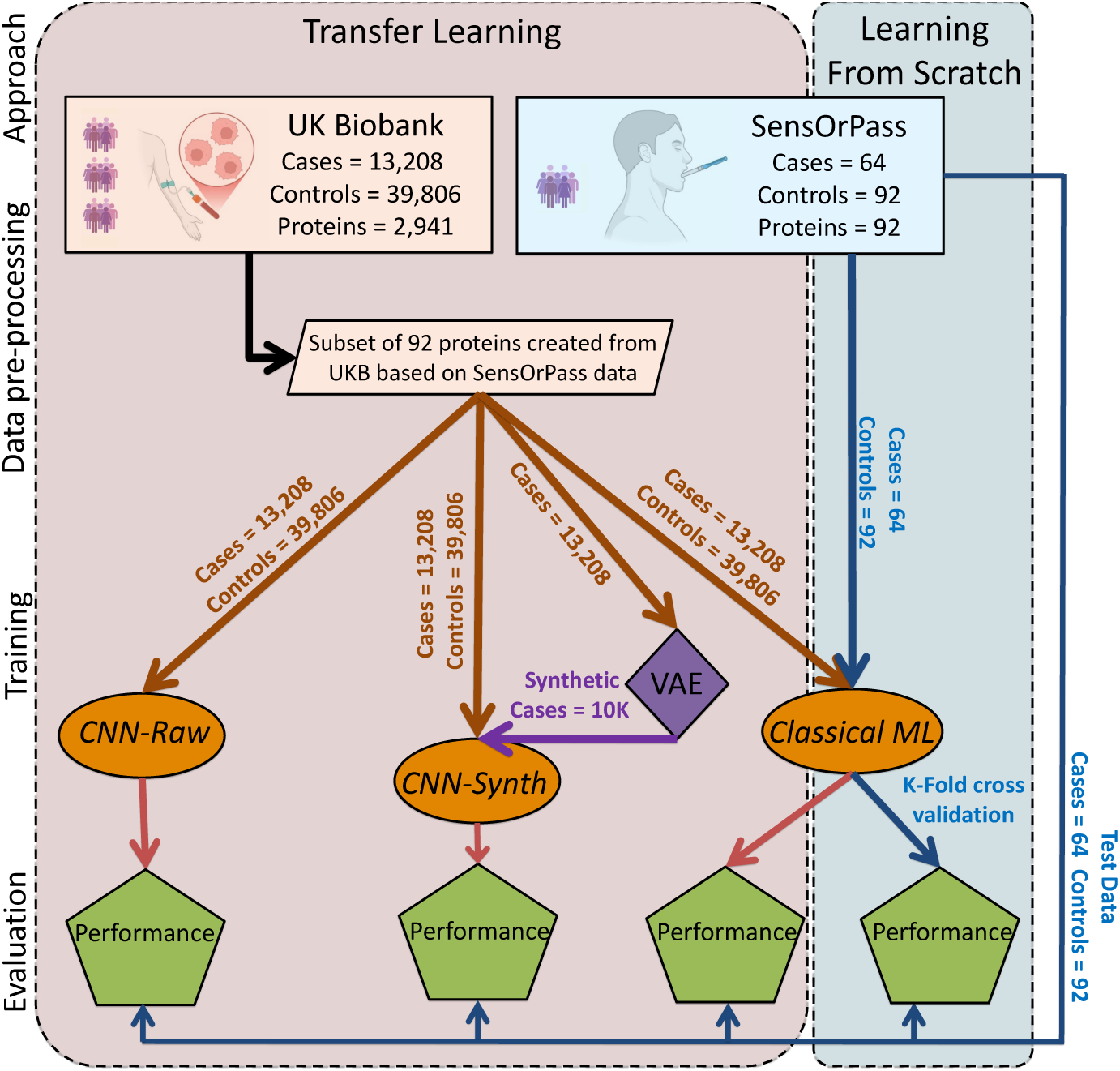
A schematic representation of the proposed pipeline. The left panel illustrates the transfer learning approach: CNN-raw & Classical ML models trained on UKB data using 92 selected protein measurements, and CNN-Synth with additional synthetic cancer cases generated by a VAE. These models are evaluated on the SensOrPass dataset. The right panel represents ‘Learning From Scratch’ where Classical-ML (e.g., LR-elasticnet, LDA, KNN, CART, NB, SVM and XGBoost) were trained and evaluated directly on the SensOrPass cohort using 10-fold cross-validation.ML: machine learning; UKB: UK Biobank; VAE: variational autoencoder; LR-elasticnet: logistic regression; LDA: linear discriminant analysis; KNN: k-nearest neighbors; CART: decision trees; NB: Gaussian naïve Bayes, SVM: support vector machines and XGBoost: eXtreme gradient boosting.

### 2.5 Classification performance

We evaluated classification performance by confusion matrices, precision (how many predicted positives are actually cancer cases), recall (how many actual cancer cases were correctly identified), and F1-score (the harmonic mean of precision and recall). We used Area Under the Receiver Operating Characteristic (ROC) curve (AUC) to summarise the classification performance for each model across the range of threshold values.

### 2.6 Model Interpretation using Shapley values

To interpret the role of individual proteins in cancer classification models, we used SHapley Additive exPlanations (SHAP) [21] within the SensOrPass dataset to estimate feature importance values. This was applied to both deep learning (CNN-Synth) and classical ML models to allow consistent comparison.

### 2.7 Implementation resources

All analyses were performed in Python versions 3.9.18. Our deep learning models, VAE (sec: 2.2) and CNN (sec: 2.3), were trained and evaluated using Keras with a TensorFlow backend (version: 2.18.0) [29]. All other machine learning models (sec: 2.4) were implemented and benchmarked for performance in scikit-learn [30]. Finally, feature importance and model explanability was performed using the SHAP Python package [21].

## 3 Results

With the aim of developing a saliva-protein classifier of head and neck cancer in SensOrPass, we trained CNN pan-cancer classification models in UKB. An overall summary of our modeling workflow and the data used is provided in Figure 1 and Table 1, respectively.

### 3.1 Training a VAE of cancer-specific protein abundance

To potentially mitigate the negative impact of class imbalance on model performance, we explored the utility of generative deep learning to synthesize case data. For this, we trained a Variational Autoencoder (VAE) on data from 13, 208 UKB cancer cases and used it to generate 10, 000 synthetic pan-cancer samples (see Methods Section 2.2). These simulated cases were added to the original training set, increasing it to 23, 208 to improve class balance (Figure 1).

The VAE effectively captured shared biological patterns between the real and synthetic data (Figure 2(b)). The latent space visualization demonstrates that the simulated pan-cancer samples occupy a similar distribution to both the original UKB cases and the SensOrPass samples. This overlap suggests that the synthetic samples are drawn from the same underlying distributions as the real data and that they are appropriate for use in augmenting imbalanced training data.

**Fig. 2.**
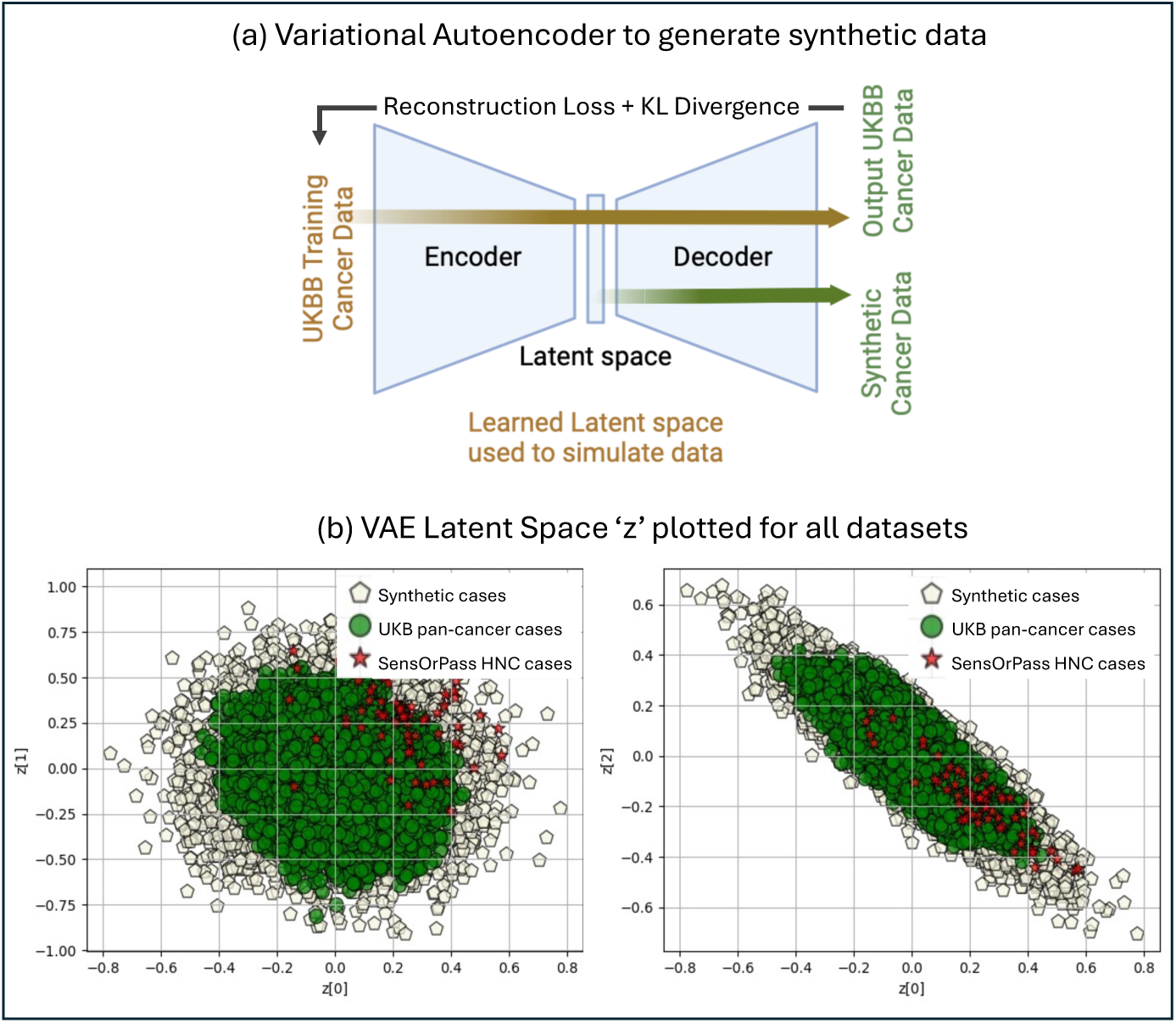
Variational Autoencoder for generation of proteomics synthetic data. **(a)** Encoder consists of four fully connected layers of size 92, 50, 25 and 10. Decoder consists of a symmetric set of four fully connected layers of size 10, 25, 50 and 92. **(b)** Latent distribution of UKB pan-cancer data, generated synthetic data and SenseOrPass HNC data. UKB: UK Biobank, HNC: head and neck cancer.

Although it would have been possible to generate sufficient synthetic samples to effectively eliminate all class imbalance, we generated only 10,000 synthetic samples to prevent overfitting. Generating too many synthetic samples increases the risk of modelling technical artefacts of synthetic data that are not present in the real-world data.

### 3.2 Classifying cancer with protein-based CNN models

We trained two CNN pan-cancer classification models in UKB (Figures 1 and 3). The first model, called CNN-Synth, was trained on data from UKB cases and controls along with 10,000 synthesized cases. The second, CNN-Raw, was trained only on UKB cases and controls to evaluate the impact of the synthesized cases on CNN-Synth. Evaluation of both models in SensOrPass revealed that the addition of synthetic samples in the CNN-Synth training dataset improved most performance metrics by a large margin (Table 2 and Figure 4(a)). Compared to CNN-Raw, there is a decrease in miss-classification both for controls and cases, false positives decreased from 17 to 11 and false negatives from 30 to 21 (Figure 4(b)).

**Fig. 3.**
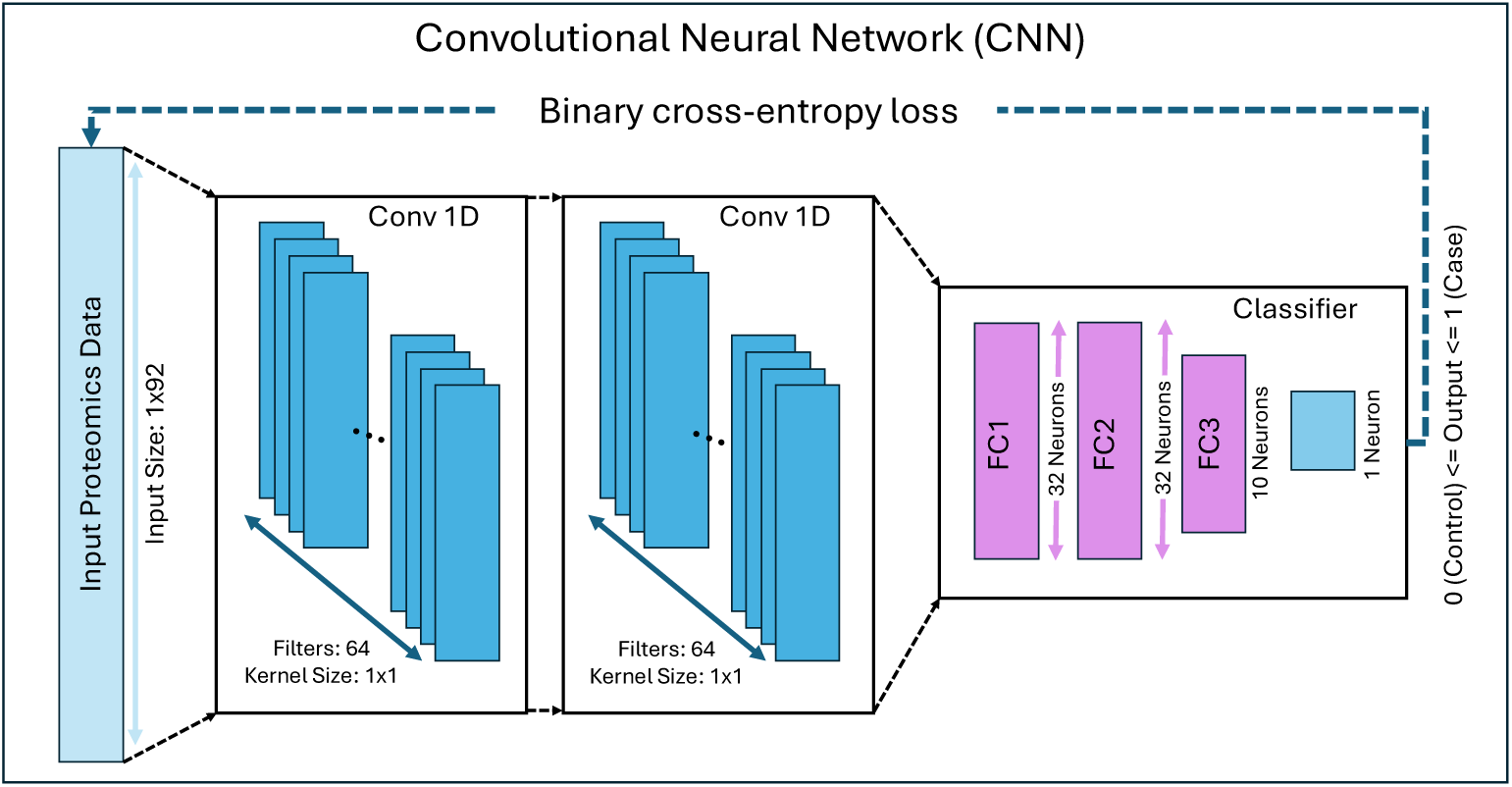
Network architecture for convolutional neural network models. Both CNN-Synth and CNN-Raw include 2 convolutional layers with 64 filters each following a classifier consisting of 3 fully connected layers of sizes 32, 32 and 10. Output consists of a single neuron with a sigmoid activation function. The model is trained using the binary cross-entropy loss function.

**Fig. 4.**
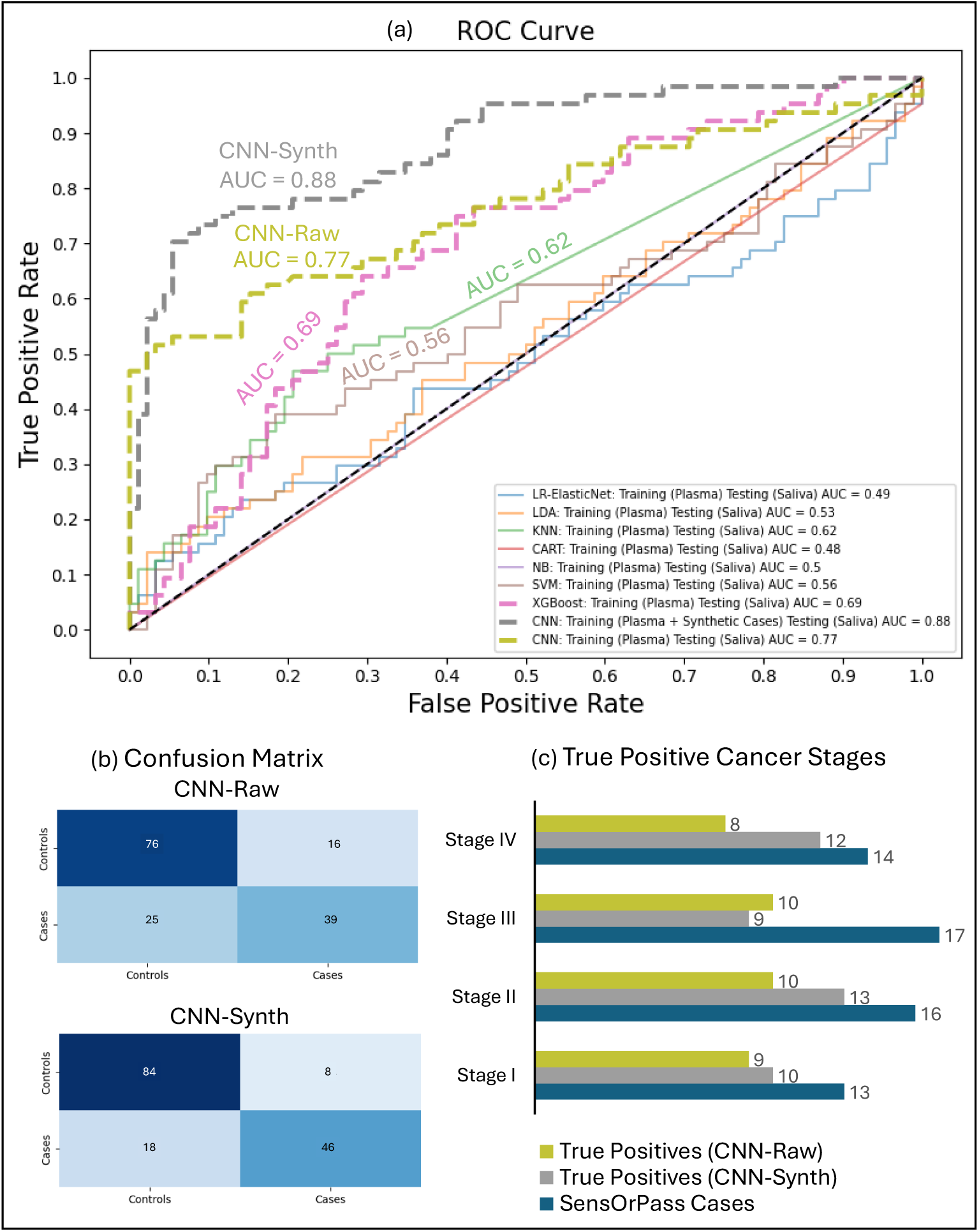
Performance evaluation of transfer learning approach. **(a)** ROC plots of all models evaluated in SensOrPass. **(b)** Confusion matrices of both CNN-Raw and CNN-Synth. **(c)** Bar plot of true positives by TNM staging for CNN-Raw (green) and CNN-Synth (grey). No information of staging was available for 4 out of 64 HNC cases in the SensOrPass study. ROC: receiver operating characteristic; AUC: area under the curve.

**Table 2.**
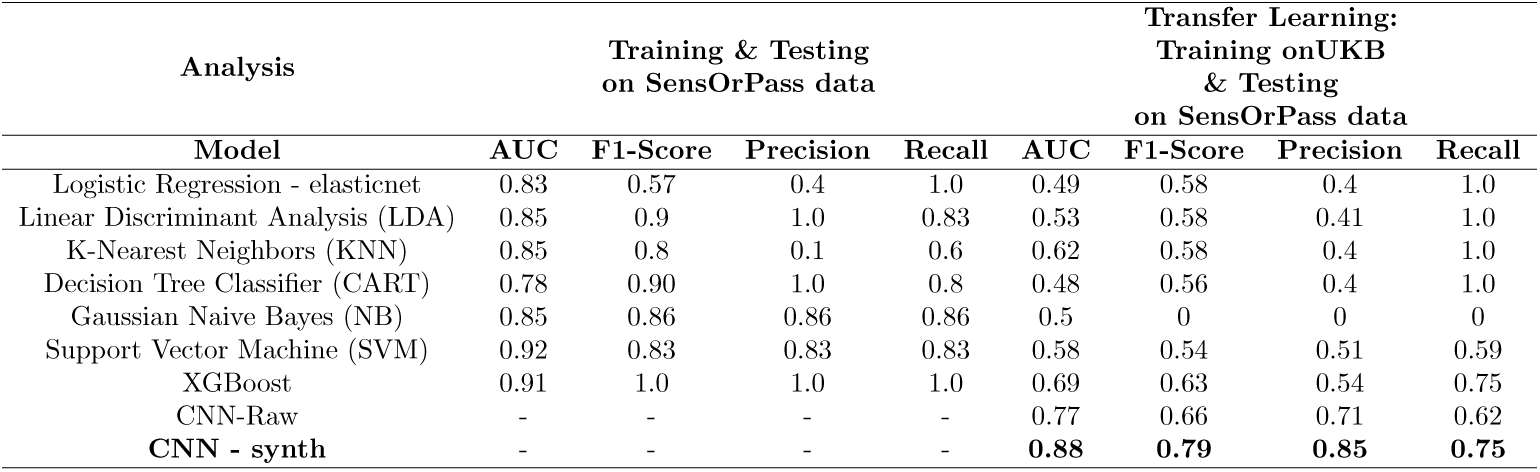
The table reports Area under the curve (AUC), F1-score and Precision and Recall for all approaches (columns) and models (rows).

To further investigate the clinical relevance of these correct predictions, we examined the cancer stage distribution of the 52 true-positive cases in CNN-Synth and 43 true positives in CNN-Raw. As shown in Figure 4(c), both CNN-Raw and CNN-Synth were able to detect cancers across a range of stages (I, II, III, IV), including early-stage disease. At all stages, CNN-Synth demonstrated greater sensitivity, and misclassification rates were highest for both models at Stage III.

### 3.3 Comparison to classical ML cancer classifiers

We compared the performance of our CNN models with a selection of classical machine learning models (see Methods Section 2.4) according to two different scenarios (Figure 1). In the transfer learning scenario (training in UKB and testing in SensOrPass - left panel shown in Figure 1), the CNN models consistently outperformed these classical machine learning models (Figure 4(a) and Table 2). After the CNN models with AUCs of 0.77 and 0.88, XGBoost has the next best performance at AUC = 0.69, followed by KNN at AUC = 0.62 and SVM at AUC=0.56.

In the training and testing scenario carried out from scratch (i.e. entirely within the SensOrPass dataset under cross-validation, right panel of Figure 1), performances were generally much higher overall (Supplementary Figure 9 (a-g)). In fact, average AUCs were as high as 0.92, slightly higher than CNN-Synth in the transfer learning scenario. However, these performances should be interpreted with caution due to the high variance in performances across the 10 cross-validation folds (Supplementary Figure 9(a) and (h)). The high variance suggests over-fitting due to small sample size. A direct comparison to a CNN model in this scenario was not made because the dataset was too small to train a CNN.

### 3.4 Model interpretation using Shapley values

The mean Shapley values in SensOrPass highlight the relative importance of salivary proteomic biomarkers for HNC detection within each trained model. We can therefore compare models by comparing their Shapley values for each protein. Figure 5(a-c) illustrates comparisons between CNN-Synth and best performing classical ML models, XGBoost, KNN and SVM. For the CNN-Synth, proteins such as IL6, CXCL17, CXCL13, IGF1R and FASLG were identified as the top five contributors, consistent with their established roles in cancer biology. Although there is some agreement with other models, correlations of Shapley values between CNN-Synth and the other models is low (Pearson’s R *<* −0.02).

**Fig. 5.**
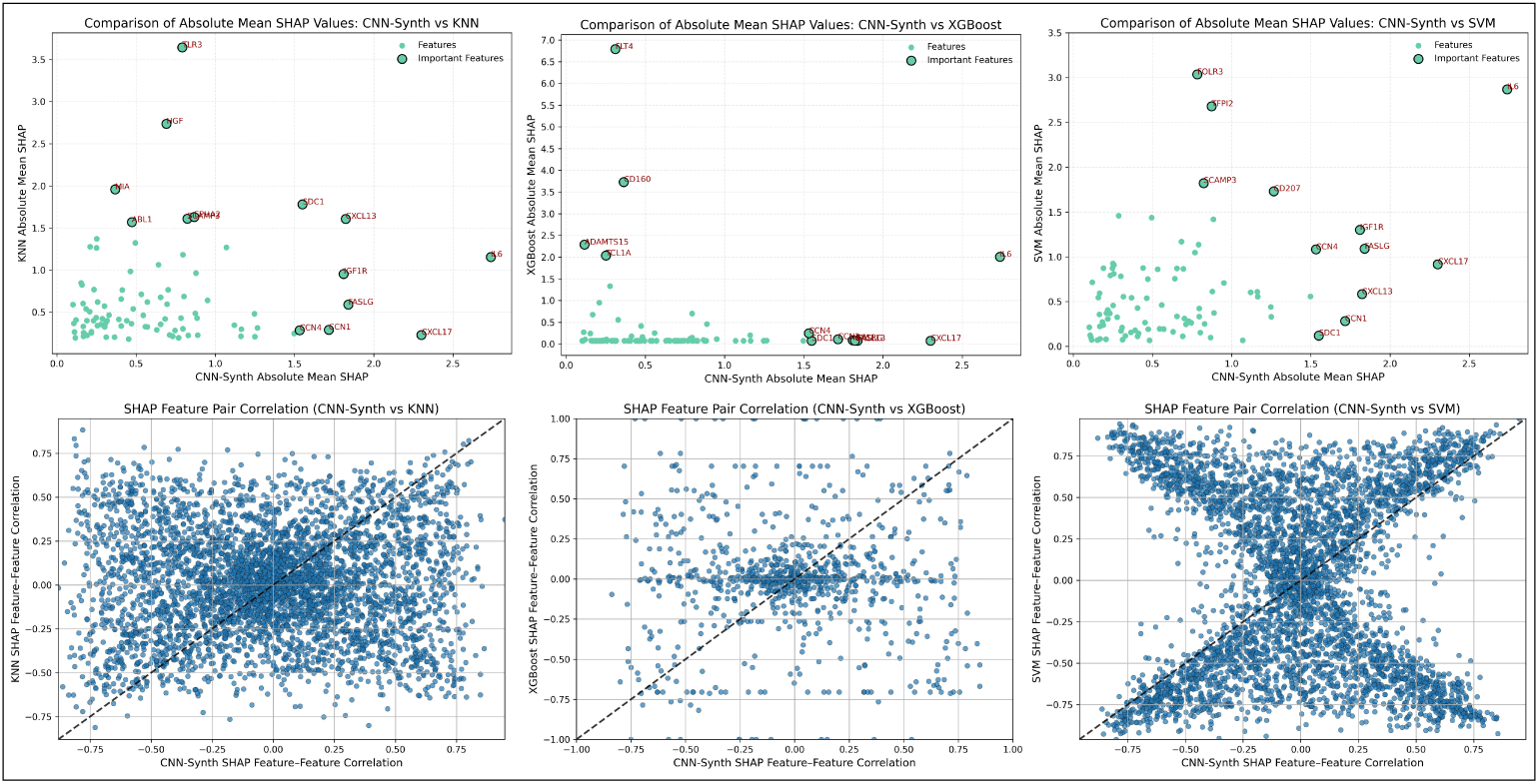
SHAP analysis for best performing models from the transfer learning approach. **(a-c)** Feature importances (mean absolute Shapley values) for CNN-Synth versus other classical ML models. **(d-e)** Pairwise feature Shapley value correlations of CNN-Synth versus those of KNN (r=0.013), XGBoost (r=-0.173) and SVM (r=-0.024). SHAP: SHapley Additive exPlanations; KNN: k-nearest neighbors; XGBoost: eXtreme gradient boosting; SVM: support vector machines.

Pair-wise correlations of Shapley values for pairs of proteins within a given model indicate the extent to which the model captures interactions between the proteins. Figure 5(d-e) compares the pair-wise correlations for CNN-Synth to the best-performing classical ML models. Overall, the interactions captured by CNN-Synth are very different from those captured by classical models, and CNN-Synth tends to capture stronger interactions between proteins, especially compared to XGBoost and SVM. KNN has a more similar range of correlations, but they are still somewhat closer to zero for KNN than for CNN-Synth.

## 4 Discussion

In this study, we investigate the ability of peripheral protein abundance to detect the presence of HNC. Given a small size typical for rare outcomes like HNC, we address this challenge using deep learning solutions based on combined application of generative models and transfer learning. Specifically, we train deep learning models in a large pan-cancer population with VAE-generated synthetic cancer protein profiles to mitigate class imbalance followed by transferring the resulting model, called CNN-Synth, to a smaller case-control study for performance evaluation. We report evidence that both transfer learning and synthetic data generation may provide independent improvements in classification performance.

Despite the success of deep learning models at solving a wide variety of general imaging and language processing problems, they have only recently been proposed for early detection of disease using omic-scale data. The requirement of large training datasets has previously prevented such applications. However, the recent generation of molecular datasets for large, population-based cohort studies like UKB has made it feasible to investigate their ability to model general molecular patterns that can be transferred to limited target datasets enriched with cases of rare diseases. We provide evidence that, indeed, deep learning with sufficient training data can outperform approaches from classical machine learning. In particular, we show that a CNN pan-cancer classifier trained in protein data derived from blood plasma generalizes well to protein data derived from a different cell type, saliva, outperforming classical machine learning models trained either from scratch or within a transfer learning context. We note that the pan-cancer training population included only about 14% with HNC.

Further, by application of generative deep learning models to measurements performed in sufficiently large sample sizes, it is feasible to model underlying distributions and interactions in protein abundance datasets that can be used to generate realistic synthetic protein profiles. Specifically, we train a VAE, a particular class of generative neural net model, on a limited number of protein measurements in a large population cohort and observe that synthetic protein abundance profiles sampled from this model correspond to the same underlying data generating distributions as the original training data. We confirmed the value of synthetic protein profiles generated by VAE by including them when training CNN-Synth. Their inclusion in training improved classification performance substantially (from AUC=0.77 to 0.88 and F1-score=0.66 to 0.79).

Our study contributes to the growing body of applied studies [11, 18] that aim to solve the class imbalance problem in omics data using generative deep learning models. [18] incorporates GANs to simulate and augment synthetic data whereas [11] uses VAE. VAEs have shown strengths in specific generative tasks that benefit from latent space representations, such as drug design [31] and dimensionality reduction of high-dimensional omics data [15]. While GANs are more commonly applied in generating realistic medical images [32] or creating synthetic EHR data for privacy preservation [33], VAEs excel in applications that require a more structured probabilistic framework for learning underlying data distributions. Like our study, both studies [11, 18] use transfer learning, but ours differs by not requiring any re-training or fine-tuning of pre-trained models in target data. For example, [11] fine-tunes the pre-trained TCGA model on each cancer subtype.

In contrast to generative approaches, traditional oversampling methods like SMOTE are still widely used but show limitations in high-dimensional biological data. For example, Safe-Level-SMOTE can amplify noise in sparse regions [34], while k-means SMOTE may overrepresent dense clusters and ignore outliers [35]. In omics settings, SMOTE often fails to reduce majority class bias and may introduce spurious correlations [36]. Our generative transfer learning approach addresses these issues by learning structured, probabilistic representations of the data, enabling more robust and generalizable modeling without relying on heuristic sample synthesis.

Approaches based on deep learning have long been criticized for not being interpretable [11, 15]. Fortunately, recent methods for understanding complex model performance have improved greatly, including SHAP [21] and LIME (Local Interpretable Model-agnostic Explanations) [37]. We selected SHAP to investigate our CNN-Synth model because it provides both global and local feature importance using Shapley values as opposed to the local-only by design behavior of LIME. Our SHAP approach prioritizes consistency and accuracy of interpretation over flexibility preferred by LIME.

Shapley values from our CNN-Synth cancer classifier highlighted roles for IL6, CXCL13, and CXCL17 as key biomarkers, adding to existing evidence supporting their potential roles in cancer pathogenesis and their utility in early cancer detection [38–40]. These findings demonstrate the potential of deep learning models for biomarker discovery and personalized cancer diagnostics, even in the context of limited data availability Comparison of Shapley values between CNN-Synth and classical machine learning models highlighted few similarities in the contribution of individual proteins to predictions. Further comparison of correlations between Shapley values for pairs of proteins aimed to illustrate the contributions of protein interactions to model predictions. Not surprisingly, we observe that a linear model like SVM utilizes little or no interaction information for prediction. XGBoost, by contrast, does appear to use some interaction information, albeit much less than the CNN model. The most flexible classical model, KNN, is most similar to CNN. However, even KNN places somewhat less importance on interactions and, for many pairs of proteins, actually reverses signs of interactions. These findings would seem to indicate that the CNN captures more complex interactions, and perhaps this improved representation of underlying interactions may be drivign its improved classification performance.

Our study has several limitations. Although our CNN performs well in data derived from saliva (SensOrPass) after having been trained data derived from blood (UKB), we suspect that performance would have been better if training and testing tissues had been the same. Our study nonetheless highlights the important and perhaps surprising overlaps in biomarkers between tissue types that were uncovered only by a deep learning model rather than simpler, more traditional machine learning models. Performance also likely suffered due to low numbers of HNC (14% of cancer cases) available in UKB for training. We therefore chose to train the model to differentiate between cases from any cancer and controls. In spite of this limitation, our study highlights important and surprising similarities in biomarkers that appear to be shared between many cancers. However, broad conclusions about similarities between cancers are limited by the fact that model testing was restricted to a single, small head and neck cancer (HNC) dataset. We hope that our study will motivate future studies to explore use of this approach for pan-cancer early detection. Our study investigated only the measurement of a small number of proteins (92). Model performances may have been improved if measurements of other proteins had been available, as well as other molecular measurements such as gene expression, DNA methylation, and/or metabolite levels. It is important to note that the 92 proteins measured in SensOrPass were drawn from a preselected panel of proteins known to be relevant to cancer diagnosis. Although this small selection may have omitted predictive proteins, it may also have ensured specificity in the CNN architecture and improved the interpretability of the resulting CNN model.

In conclusion, our results suggest that integration of deep learning with transfer learning from large-scale plasma datasets and synthetic sample generation can overcome challenges in disease detection associated with limited sample size, tissue differences and class imbalance. This approach sets a foundation for future work focused on improving early stage classification and expanding clinical applicability that are often faced with similar challenges.

## 5 Data Sharing Statement

UKB data are available to access by application procedure detailed here: http://www.ukbiobank.ac.uk/using-the-resource/. SensOrPass data, study protocol, and data dictionary are available from the Head and Neck 5000 and Exeter 10,000 research resources. Full application details are available here: https://headandneck5000.org.uk/information-for-researchers/ https://exetercrfnihr.org/about/exeter-10000/.

## Data Availability

UKB data are available to access by application procedure detailed here: http://www.ukbiobank.ac.uk/using-the-resource/. SensOrPass data, study protocol, and data dictionary are available from the Head and Neck 5000 and Exeter 10,000 research resources. Full application details are available here: https://headandneck5000.org.uk/information-forresearchers/ https://exetercrfnihr.org/about/exeter-10000/

https://headandneck5000.org.uk/information-for-researchers/

http://www.ukbiobank.ac.uk/using-the-resource/

## Acknowledgment

We sincerely appreciate all UKB participants, staff, and research team members for their past and ongoing contributions to these studies. This research has been conducted using the UK Biobank Resource under Application ID 236527. The UK Medical Research Council and Wellcome (Grant ref: 217065/Z/19/Z) and the University of Bristol provide core support for the data. This publication is the work of the authors and they will serve as guarantors for the contents of this paper. P.Y. and M.S. work is supported by the National Institute for Health and Care Research Bristol Biomedical Research Centre, the Medical Research Council Integrative Epidemiology Unit at the University of Bristol (MC UU 00032\3, MC UU 00032\4, MC UU 00032\6), and Cancer Research UK [C18281\A29019, EDDISA-Jan22\100003]. S.W.D.M is supported by the National Institute for Health and Care Research (NIHR) Manchester Biomedical Research Centre (BRC) (NIHR203308). For the purpose of open access, the author has applied a CC BY public copyright license to any Author Accepted Manuscript version arising from this submission.

## 7 Author’s Contribution

S.W.D.M, A.S.M, Z.S.A and P.Y secured funding for the SensOrPass study. S.W.D.M was the PI on the SensOrPass clinical study and was responsible for data collection. J.S. was the local PI at RDUH Exeter and contributed to the recruitment of head and neck cancer patients. A.S., P.Y., M.S. and Z.S.A designed the study. A.S. and P.Y. obtained and managed access to UK Biobank data. A.S. conducted all analyses, interpreted the findings, and drafted the manuscript. P.Y., M.S., Z.S.A., S.W.D.M., and A.S.M. contributed to the drafting and critical revision of the manuscript.

## Supplemental Material

**Table 3.** UKB cohort details.

|  |  | Missing | Overall | Case | Control |
| --- | --- | --- | --- | --- | --- |
| n |  |  | 53,014 | 13,208 | 39,806 |
| Age, mean (SD) |  | 0 | 56.8 (8.2) | 59.9 (7.2) | 55.8 (8.3) |
| Gender, n (%) | Female |  | 28,580 (53.9) | 7,050 (53.4) | 21,530 (54.1) |
|  | Male |  | 24,434 (46.1) | 6,158 (46.6) | 18,276 (45.9) |
| Smoking status, n (%) | Current |  | 5600 (10.6) | 1,475 (11.2) | 4,125 (10.4) |
|  | Never |  | 28,670 (54.1) | 6,430 (48.7) | 22,240 (55.9) |
|  | None |  | 61 (0.1) | 10 (0.1) | 51 (0.1) |
|  | Prefer not to answer |  | 194 (0.4) | 51 (0.4) | 143 (0.4) |
|  | Previous |  | 18,489 (34.9) | 5,242 (39.7) | 13,247 (33.3) |
| Alcohol status, n (%) | Current |  | 48,306 (91.1) | 12,132 (91.9) | 36,174 (90.9) |
|  | Never |  | 2506 (4.7) | 512 (3.9) | 1,994 (5.0) |
|  | None |  | 61 (0.1) | 10 (0.1) | 51 (0.1) |
|  | Prefer not to answer |  | 80 (0.2) | 18 (0.1) | 62 (0.2) |
|  | Previous |  | 2,061 (3.9) | 536 (4.1) | 1,525 (3.8) |

**Table 4.** SensOrPass cohort details.

|  |  | Missing | Overall | HNC Case | Control |
| --- | --- | --- | --- | --- | --- |
| n |  |  | 156 | 64 | 92 |
| Age, mean (SD) |  | 0 | 63.6 (9.7) | 61.0 (8.5) | 65.3 (10.2) |
| Family history of cancer, n (%) | No |  | 22 (14.1) | 5 (7.8) | 17 (18.5) |
|  | Yes |  | 79 (50.6) | 7 (10.9) | 72 (78.3) |
|  | Don't know |  | 5 (3.2) | 2 (3.1) | 3 (3.3) |
|  | None |  | 50 (32.1) | 50 (78.1) |  |
| Gender, n (%) | Male |  | 89 (57.1) | 54 (84.4) | 35 (38.0) |
|  | Female |  | 67 (42.9) | 10 (15.6) | 57 (62.0) |
| Smoking status, n (%) | Yes-cigarettes |  | 8 (5.1) | 7 (10.9) | 1 (1.1) |
|  | Ex-smoker |  | 70 (44.9) | 39 (60.9) | 31 (33.7) |
|  | Never smoked |  | 78 (50.0) | 18 (28.1) | 60 (65.2) |
| Alcohol status, n (%) | No |  | 23 (14.7) | 7 (10.9) | 16 (17.4) |
|  | Yes |  | 133 (85.3) | 57 (89.1) | 76 (82.6) |
| BMI, mean (SD) |  | 0 | 27.0 (4.8) | 27.1 (4.6) | 26.8 (4.8) |

**Fig. 6.**
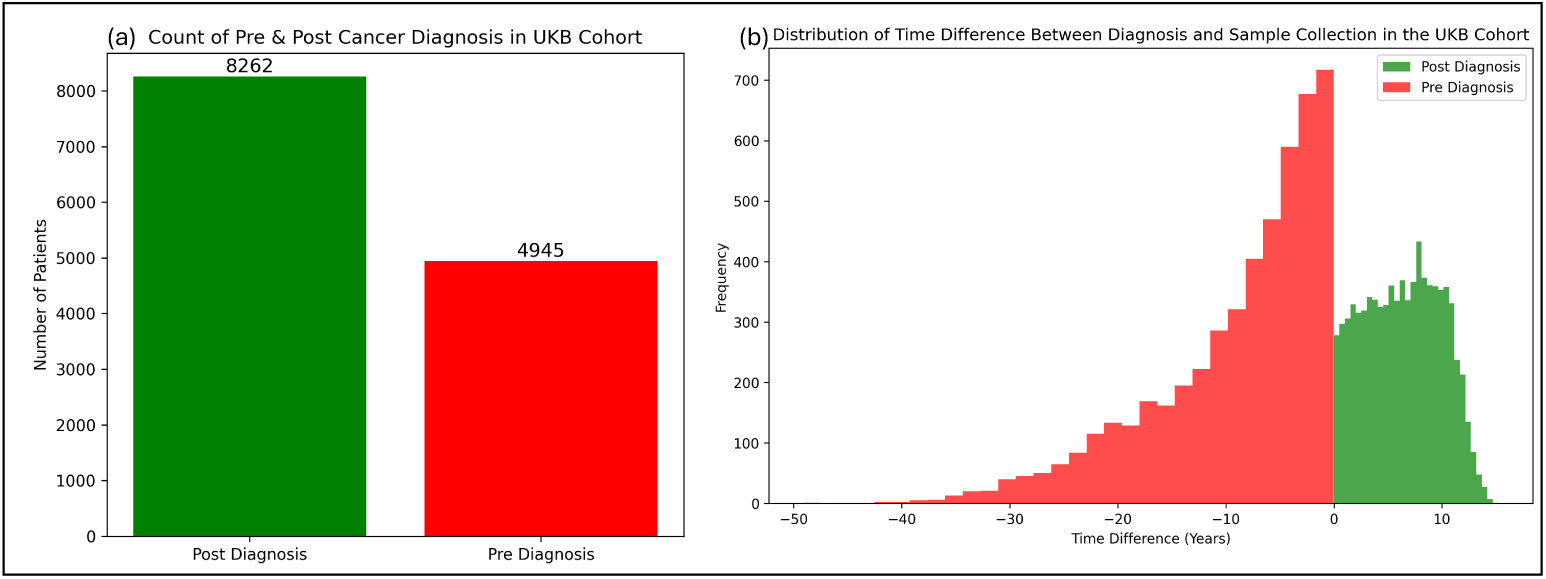
Timing of cancer diagnosis relative to sample collection in UKB. **(a)** Bar plot of cancer diagnosis pre and post sample collection in the UKB data cohort. **(b)** Distribution of change in time when the cancer was first diagnosed and when the samples where drawn for UKB participants. UKB: UK Biobank.

**Fig. 7.**
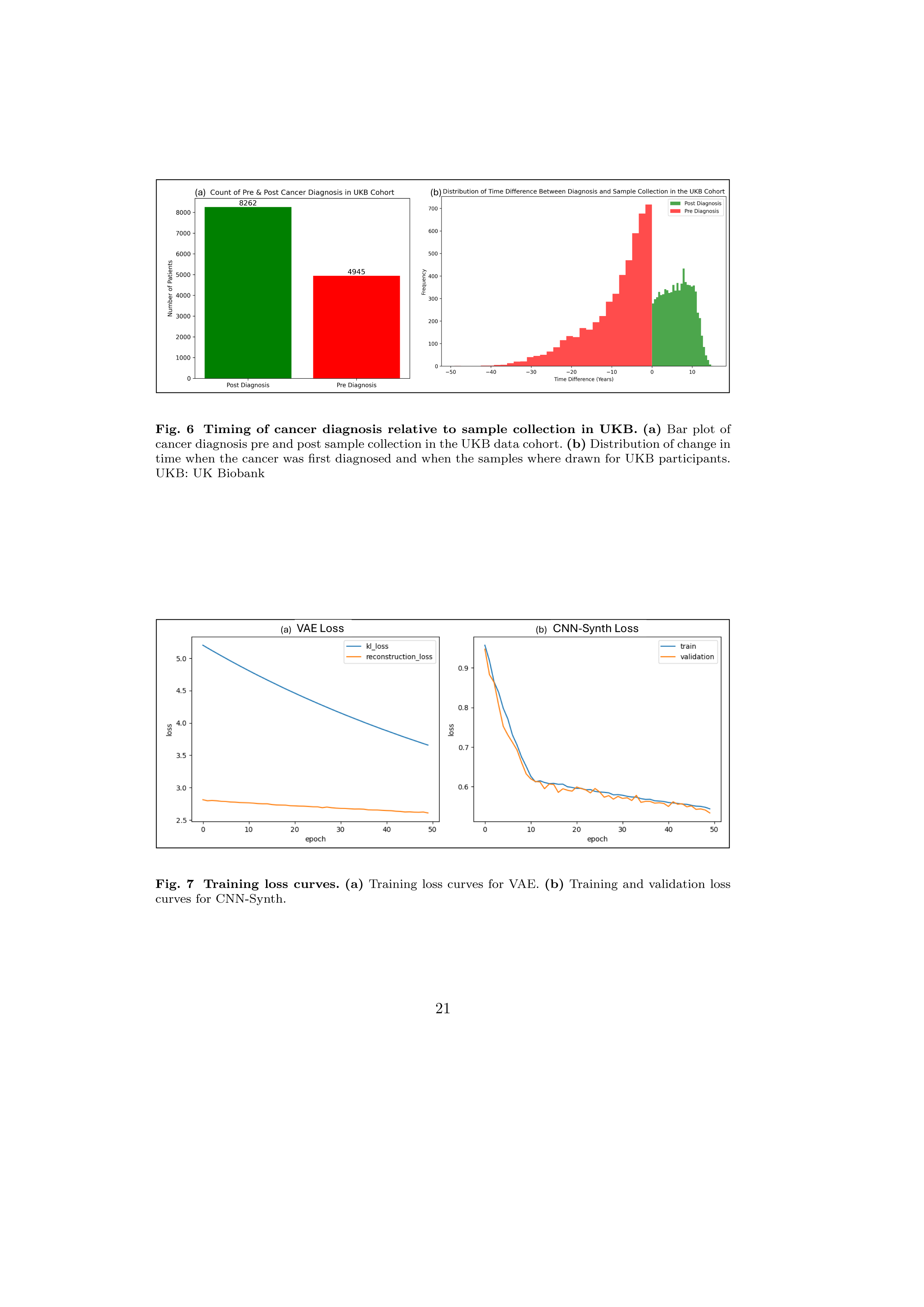
Training loss curves. **(a)** Training loss curves for VAE. **(b)** Training and validation loss curves for CNN-Synth.

**Fig. 8.**
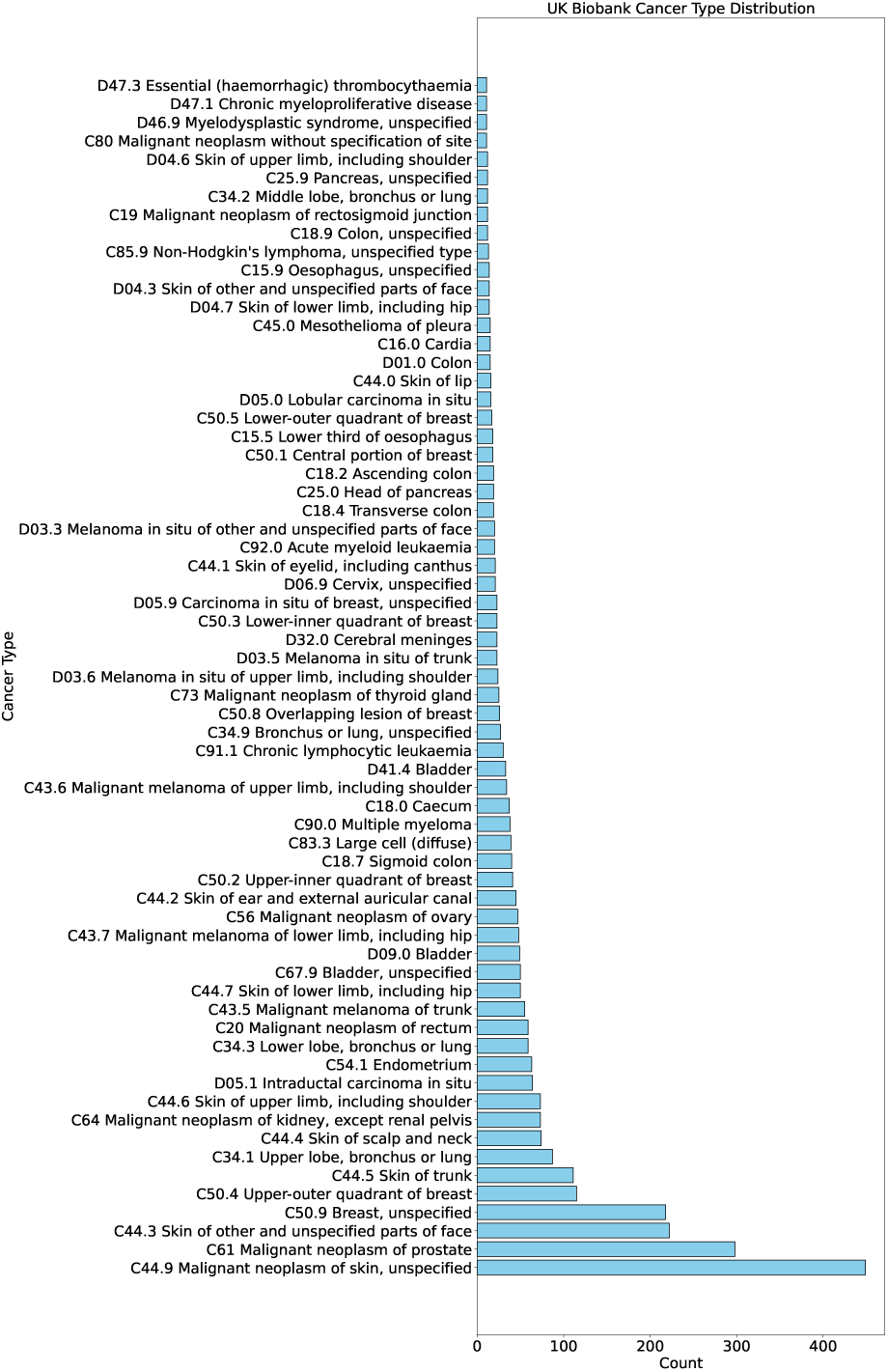
Types of cancer recorded in the UKB dataset.

**Fig. 9.**
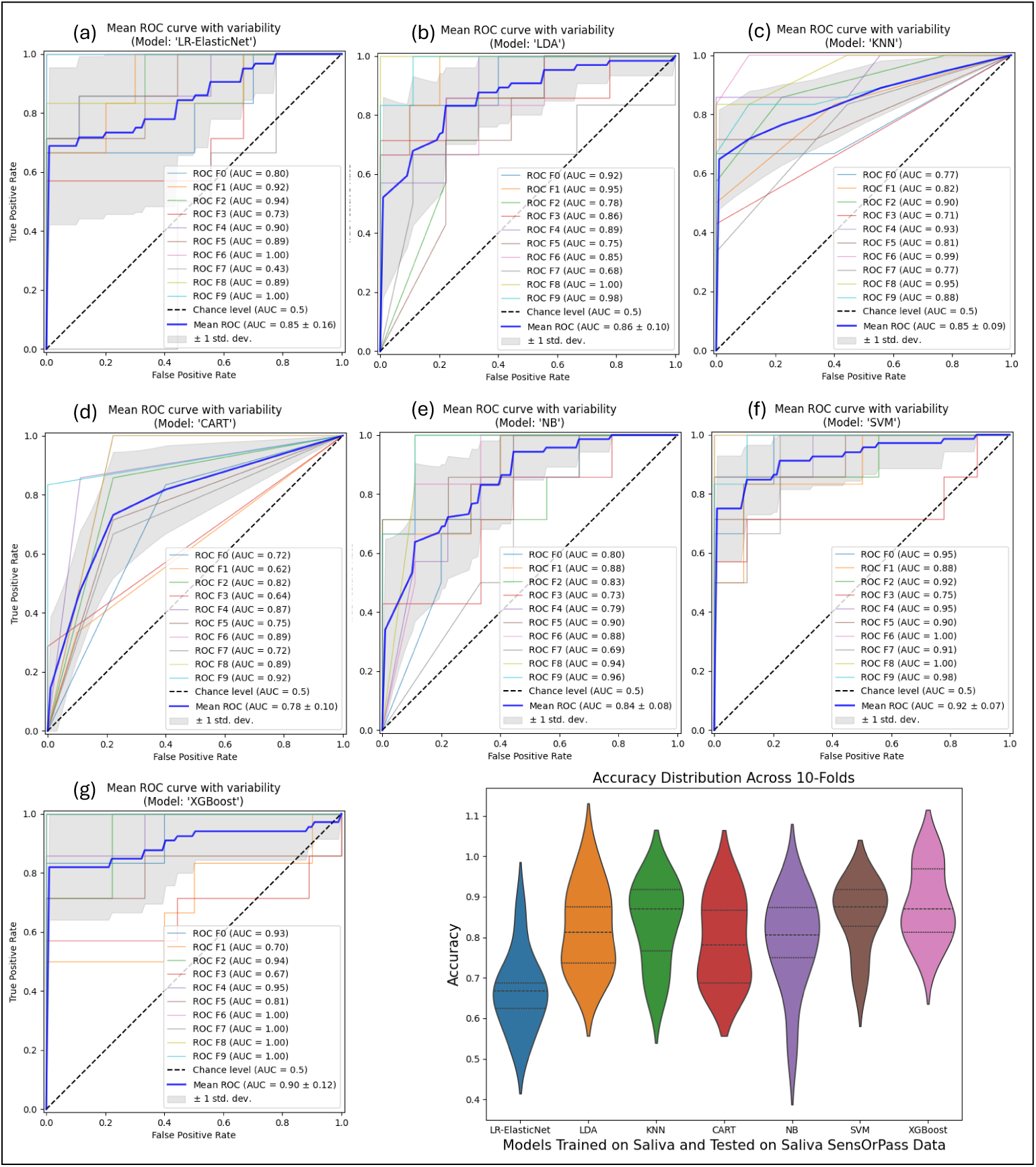
Model Performance When Trained from Scratch. **(a-g)** Shows the ROC plot of supervised machine learning models that are trained and tested on saliva proteome data from SensOrPass study. **(b)** Shows the variance of accuracy across the 10 folds of cross-validation for each of these machine learning models.

## References

[1] Gormley, M., Creaney, G., Schache, A., Ingarfield, K., Conway, D.I.: Reviewing the epidemiology of head and neck cancer: definitions, trends and risk factors. British Dental Journal 233(9), 780–786 (2022)

[2] Chow, L.Q.: Head and neck cancer. New England Journal of Medicine 382(1), 60–72 (2020)

[3] Carrasco-Zanini, J., Pietzner, M., Davitte, J., Surendran, P., Croteau-Chonka, D.C., Robins, C., Torralbo, A., Tomlinson, C., Grünschläger, F., Fitzpatrick, N., et al.: Proteomic signatures improve risk prediction for common and rare diseases. Nature medicine 30(9), 2489–2498 (2024)

[4] Gadd, D.A., Hillary, R.F., Kuncheva, Z., Mangelis, T., Cheng, Y., Dissanayake, M., Admanit, R., Gagnon, J., Lin, T., Ferber, K., et al.: Blood protein levels predict leading incident diseases and mortality in uk biobank. medRxiv, 2023–05 (2023)

[5] Budach, V., Tinhofer, I.: Novel prognostic clinical factors and biomarkers for outcome prediction in head and neck cancer: a systematic review. The Lancet Oncology 20(6), 313–326 (2019)

[6] Dikova, V., Jantus-Lewintre, E., Bagan, J.: Potential non-invasive biomarkers for early diagnosis of oral squamous cell carcinoma. Journal of Clinical Medicine 10(8), 1658 (2021)

7. The blood proteome of imminent lung cancer diagnosis. Nature communications 14(1), 3042 (2023)

[8] Sun, B.B., Chiou, J., Traylor, M., Benner, C., Hsu, Y.-H., Richardson, T.G., Surendran, P., Mahajan, A., Robins, C., Vasquez-Grinnell, S.G., et al.: Plasma proteomic associations with genetics and health in the uk biobank. Nature 622(7982), 329–338 (2023)

[9] Topol, E.J.: High-performance medicine: the convergence of human and artificial intelligence. Nature medicine 25(1), 44–56 (2019)

[10] Esteva, A., Kuprel, B., Novoa, R.A., Ko, J., Swetter, S.M., Blau, H.M., Thrun, S.: Dermatologist-level classification of skin cancer with deep neural networks. nature 542(7639), 115–118 (2017)

[11] Karlberg, B., Kirchgaessner, R., Lee, J., Peterkort, M., Beckman, L., Goecks, J., Ellrott, K.: Synthevaeiser: augmenting traditional machine learning methods with vae-based gene expression sample generation for improved cancer subtype predictions. Genome biology 25(1), 1–18 (2024)

[12] LeCun, Y., Bengio, Y., Hinton, G.: Deep learning. nature 521(7553), 436–444 (2015)

[13] Ballard, J.L., Wang, Z., Li, W., Shen, L., Long, Q.: Deep learning-based approaches for multi-omics data integration and analysis. BioData Mining 17(1), 38 (2024)

[14] Zompola, A., Korfiati, A., Theofilatos, K., Mavroudi, S. OMICS-CNN: A comprehensive pipeline for predictive analytics in quantitative omics using one dimensional convolutional neural networks (2023) 10.2139/ssrn.4427000

15. Zhuang, J., Smith, E.N., Diogo, D.: Representation learning based on proteomic profiles uncovers key cell types and biological processes contributing to the plasma proteome. medRxiv (2024) 10.1101/2024.12.16.24319106 https://www.medrxiv.org/content/early/2024/12/16/2024.12.16.24319106.full.pdf

[16] Bozinovski, S., Fulgosi, A.: The influence of pattern similarity and transfer learning upon training of a base perceptron b2. In: Proceedings of Symposium Informatica, vol. 3, pp. 121–126 (1976)

[17] Libbrecht, M.W., Noble, W.S.: Machine learning applications in genetics and genomics. Nature Reviews Genetics 16(6), 321–332 (2015)

[18] Cusworth, S., Gkoutos, G.V., Acharjee, A.: A novel generative adversarial networks modelling for the class imbalance problem in high dimensional omics data. BMC Medical Informatics and Decision Making 24(1), 90 (2024)

[19] Nicholas, I., Kuo, H., Garcia, F., Sonnerborg, A., Bohm, M., Kaiser, R., Zazzi, M., Polizzotto, M., Jorm, L., Barbieri, S., et al.: Generating synthetic clinical data that capture class imbalanced distributions with generative adversarial networks: example using antiretroviral therapy for hiv. Journal of Biomedical Informatics 144, 104436 (2023)

[20] Hira, M.T., Razzaque, M.A., Angione, C., Scrivens, J., Sawan, S., Sarker, M.: Integrated multi-omics analysis of ovarian cancer using variational autoencoders. Scientific reports 11(1), 6265 (2021)

[21] Lundberg, S.: A unified approach to interpreting model predictions. arXiv preprint arXiv:1705.07874 (2017)

[22] Wik, L., Nordberg, N., Broberg, J., Bjorkesten, J., Assarsson, E., Henriksson, S., Grundberg, I., Pettersson, E., Westerberg, C., Liljeroth, E., et al.: Proximity extension assay in combination with next-generation sequencing for high-throughput proteome-wide analysis. Molecular & Cellular Proteomics 20 (2021)

[23] Ness, A.R., Waylen, A., Hurley, K., Jeffreys, M., Penfold, C., Pring, M., Leary, S., Allmark, C., Toms, S., Ring, S., et al.: Establishing a large prospective clinical cohort in people with head and neck cancer as a biomedical resource: head and neck 5000. Bmc Cancer 14, 1–6 (2014)

[24] Ness, A.R., Waylen, A., Hurley, K., Jeffreys, M., Penfold, C., Pring, M., Leary, S., Allmark, C., Toms, S., Ring, S., et al.: Recruitment, response rates and characteristics of 5511 people enrolled in a prospective clinical cohort study: head and neck 5000. Clinical Otolaryngology 41(6), 804 (2016)

[25] Exeter CRF: Exeter Clinical Research Facility // Exeter 10,000 — exetercrfnihr.org. https://exetercrfnihr.org/about/exeter-10000/. [Accessed 26-02-2025]

[26] Wang, S., Wang, S., Wang, Z.: A survey on multi-omics-based cancer diagnosis using machine learning with the potential application in gastrointestinal cancer. Frontiers in Medicine 9, 1109365 (2023)

[27] Yamashita, R., Nishio, M., Do, R.K.G., Togashi, K.: Convolutional neural networks: an overview and application in radiology. Insights into imaging 9, 611–629 (2018)

[28] Liao, J., Li, X., Gan, Y., Han, S., Rong, P., Wang, W., Li, W., Zhou, L.: Artificial intelligence assists precision medicine in cancer treatment. Frontiers in oncology 12, 998222 (2023)

29. Abadi, M., Agarwal, A., Barham, P., Brevdo, E., Chen, Z., Citro, C., Corrado, G.S., Davis, A., Dean, J., Devin, M., et al.: Tensorflow: Large-scale machine learning on heterogeneous distributed systems. arXiv preprint arXiv:1603.04467 (2016)

[30] Pedregosa, F., Varoquaux, G., Gramfort, A., Michel, V., Thirion, B., Grisel, O., Blondel, M., Prettenhofer, P., Weiss, R., Dubourg, V., et al.: Scikit-learn: Machine learning in python. the Journal of machine Learning research 12, 2825–2830 (2011)

[31] Gomez-Bombarelli, R., Wei, J.N., Duvenaud, D., Herńandez-Lobato, J.M., Śanchez-Lengeling, B., Sheberla, D., Aguilera-Iparraguirre, J., Hirzel, T.D., Adams, R.P., Aspuru-Guzik, A.: Automatic chemical design using a data-driven continuous representation of molecules. ACS central science 4(2), 268–276 (2018)

32. Bowles, C., Chen, L., Guerrero, R., Bentley, P., Gunn, R., Hammers, A., Dickie, D.A., Herńandez, M.V., Wardlaw, J., Rueckert, D.: Gan augmentation: Augmenting training data using generative adversarial networks. arXiv preprint arXiv:1810.10863 (2018)

33. Beaulieu-Jones, B.K., Wu, Z.S., Williams, C., Lee, R., Bhavnani, S.P., Byrd, J.B., Greene, C.S.: Privacy-preserving generative deep neural networks support clinical data sharing. Circulation: Cardiovascular Quality and Outcomes 12(7), 005122 (2019)

[34] Bunkhumpornpat, C., Sinapiromsaran, K., Lursinsap, C.: Safe-level-smote: Safe-level-synthetic minority over-sampling technique for handling the class imbalanced problem. In: Advances in Knowledge Discovery and Data Mining: 13th Pacific-Asia Conference, PAKDD 2009 Bangkok, Thailand, April 27-30, 2009 Proceedings 13, pp. 475–482 (2009). Springer

[35] Douzas, G., Bacao, F., Last, F.: Improving imbalanced learning through a heuristic oversampling method based on k-means and smote. Information sciences 465, 1–20 (2018)

[36] Blagus, R., Lusa, L.: Smote for high-dimensional class-imbalanced data. BMC bioinformatics 14, 1–16 (2013)

[37] Ribeiro, M.T., Singh, S., Guestrin, C.: “why should i trust you?” explaining the predictions of any classifier. In: Proceedings of the 22nd ACM SIGKDD International Conference on Knowledge Discovery and Data Mining, pp. 1135–1144 (2016)

[38] Grivennikov, S.I., Greten, F.R., Karin, M.: Immunity, inflammation, and cancer. Cell 140(6), 883–899 (2010)

[39] Duffy, S.A., Taylor, J.M., Terrell, J.E., Islam, M., Li, Y., Fowler, K.E., Wolf, G.T., Teknos, T.N.: Interleukin-6 predicts recurrence and survival among head and neck cancer patients. Cancer: Interdisciplinary International Journal of the American Cancer Society 113(4), 750–757 (2008)

[40] Razis, E., Kalogeras, K.T., Kotsantis, I., Koliou, G.-A., Manousou, K., Wirtz, R., Veltrup, E., Patsea, H., Poulakaki, N., Dionysopoulos, D., et al.: The role of cxcl13 and cxcl9 in early breast cancer. Clinical Breast Cancer 20(1), 36–53 (2020)

